# Spatiotemporal patterns and progression of the Delta variant of COVID-19 and their health intervention linkages in Southeast Asia

**DOI:** 10.1101/2021.12.20.21268140

**Authors:** Luo Wei, Liu Zhaoyin, Zhou Yuxuan, Zhao Yumin, Li Yunyue Elita, Arif Masrur, Manzhu Yu

**Affiliations:** Department of Geography, National University of Singapore, Singapore; Department of Civil and Environmental Engineering, National University of Singapore, Singapore; Department of Earth, Atmospheric, and Planetary Sciences, Purdue University, West Lafayette, Indiana, U.S.A.; Department of Geography, Penn State University, U.S.A.

**Keywords:** Covid-19, Delta variant, Space-time Scan, Intervention, Southeast Asia

## Abstract

The global pandemic of COVID-19 presented an unprecedented challenge to all countries in the world, among which Southeast Asia (SEA) countries managed to maintain and mitigate the first wave of COVID-19 in 2020. However, these countries were caught in the crisis after the Delta variant was introduced to SEA, though many countries had immediately implemented non-pharmaceutical intervention (NPI) measures along with vaccination in order to contain the disease spread. To investigate the potential linkages between epidemic dynamics and public health interventions, we adopted a prospective space-time scan method to conduct spatiotemporal analysis at the district level in the seven selected countries in SEA from June 2021 to October 2021. Results reveal the spatial and temporal propagation and progression of COVID-19 risks relative to public health measures implemented by different countries. Our research benefits continuous improvements of public health strategies in preventing and containing this pandemic.

## 1. Introduction

Coronavirus disease 2019 (COVID-19) is a global epidemic caused by severe acute respiratory syndrome coronavirus 2 (SARS-CoV-2). SARS-CoV-2 is highly contagious as it easily spreads across human, animals and the environment via contact, droplet, airborne, fomite and other transmission modes [1]. In December 2019, COVID-19 was first identified in Wuhan – a transportation and communication hub located in central China, and rapidly spread to surrounding regions in China. Although a certain level of precautions and control regulation were taken by governments, international cases managed to emerge along with significant outbreaks worldwide. As of December 12, 2021, the disease had infected over two hundred million people across all continents except from Antarctica [2].

With the outbreak of COVID-19, countries in SEA met unprecedented challenges. Health and social care system, as well as tourism, trade in essential goods and services of SEA countries were under pressure [3]. Hence, SEA countries with export-oriented economies like Cambodia, Vietnam, Singapore etc. suffered varying degrees of damage due to COVID-19 [3, 4]. Many SEA countries had made efforts to recover the economy during the late 2020 and early 2021 by progressively easing lockdown and strengthening export orders, but they were continuously hammered by the new wave of COVID-19 delta variant [5]. The Delta variant is estimated to be twice or four times more transmissible than its original virus with a Reproductive number (R0) as nearly 7 [6]. Since April 2021, the Delta variant of SARS-CoV-2 has caused an exponential increase of new cases in SEA that has become an emerging hotspot region of COVID-19 [7]. The new outbreak caused by delta variant heavily disrupted their national and international business due to interruption of supply chain of many productions (e.g., garment, auto parts, semiconductors) which had great impacts on the manufacture industries in SEA, especially for those relying on them (e.g., Vietnam. Malaysia, Thailand, Indonesia, and Philippines) [5]. Due to the economic pressure caused by the epidemic, a growing number of Southeast Asian countries were planning to live with the virus, along with adjusting their public health intervention policies. [8]. Under this circumstance, monitoring outbreaks and identifying the space-time clusters of infection have become significant for a coordinated response to the epidemic in SEA.

Spatio-temporal analysis has been widely used in research of COVID-19 propagation to illustrate the characteristics and mechanism of COVID-19 spatial propagation, which provided public health authorities with important information to help mitigate the situation [9-11]. Among diverse spatio-temporal methods, space-time scan is one of the most popular methods adopted to explore spatiotemporal clusters in a particular region around the world, such as Mainland China [12], the United States [13], Mexico [14], Spain [15], Malaysia [16], Bangladesh [17] etc. In SEA, previous studies have applied this analysis to investigate the first wave of COVID-19 cases in SEA [18, 19]. However, those studies mainly focused on each individual country without exploring propagation patterns and progression characteristics with collective public health interventions at the regional scale. Previous research has shown that regional-wide coordination could interrupt transmission of COVID-19 in an effective way [10, 17, 20-22]. In order to contain the emerging epidemic of COVID-19 and minimize its risk, countries in SEA have deployed various preventive and containment measures such as lockdowns, distancing restrictions, and mandatory tracking and trace method [23, 24].

Delta variant transmission had shown significant spatial heterogeneity in SEA because different countries adopted different interventions along with the spread. Hence, this paper aims to identify the space-time clusters of outbreaks of COVID-19 caused by SARS-CoV-2 delta variant in SEA. We utilized district-scale daily confirmed cases of seven SEA countries from June 2021 to October 2021 to identify the active and emerging clusters of the disease outbreaks and summarized the relative policies to investigate the potential linkage between government interventions and the transmission. Our work contributes to regional surveillance of COVID-19 progression in SEA and provides essential information of COVID-19 propagation from space and time perspective to public health authorities, which is beneficial to timely policy making according to the dynamic COVID-19 situation.

## 2. Materials and Methods

### 2.1. Study areas and relevant interventions

Our study focused on seven countries in SEA, namely Indonesia, Malaysia, the Philippines, Singapore, Thailand, Vietnam and Brunei, because they disclose relevant data at a district level. During the second COVID-19 outbreak caused by the Delta variant, the dynamic of interventions implemented by different countries along the way may cause the fluctuation of transmission. For example, Thailand, Singapore, Malaysia, Philippines, and Vietnam began to relax their restrictions around August, which may cause significant changes in the epidemic patterns after then (Table 1). The diverse policies will help explain the progression and transmission of the Delta variant of COVID-19 in the following analysis. The interventions are aggregated by Center for Strategic & International Studies (CSIS) [25].

**Table 1.**
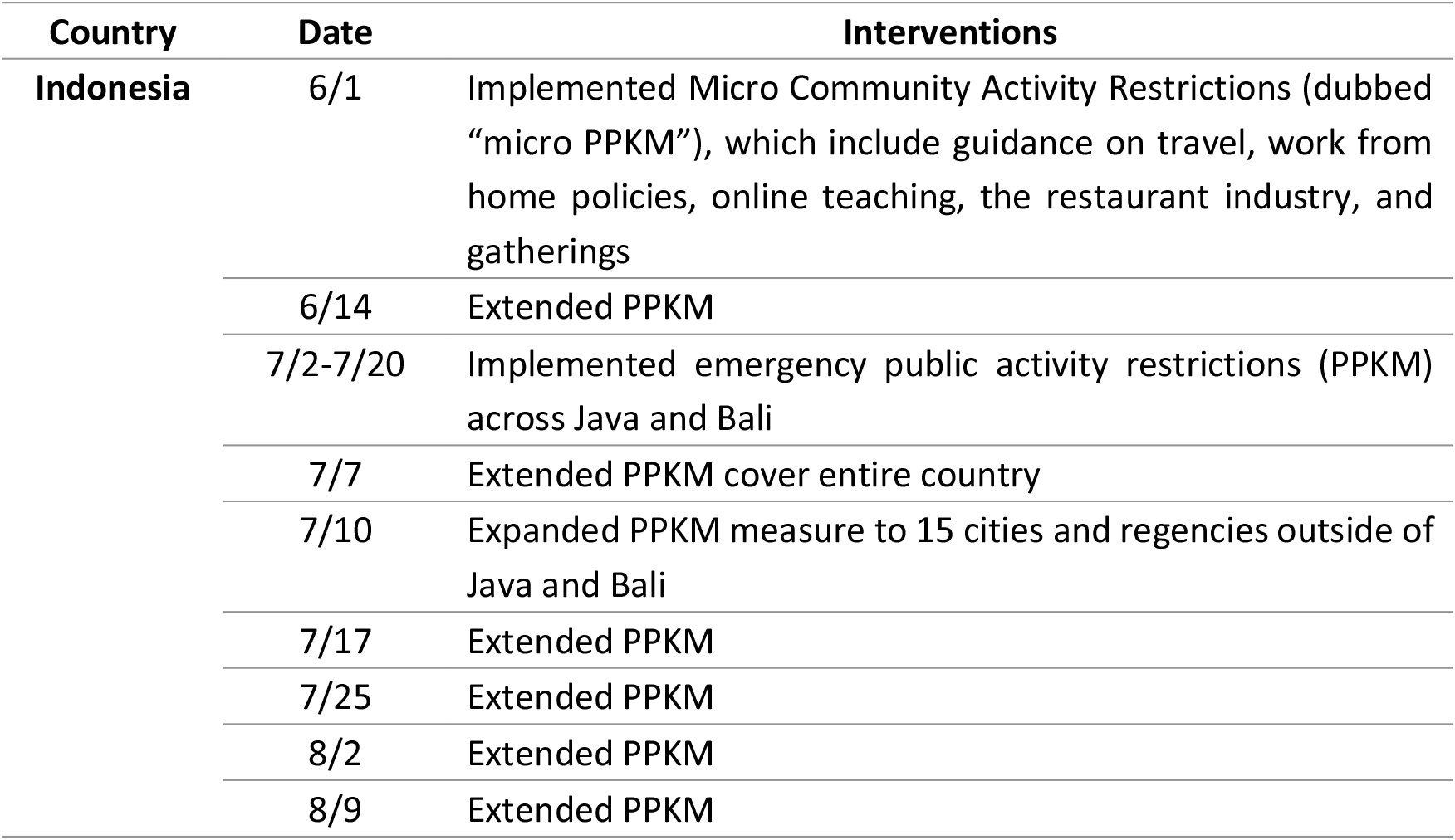
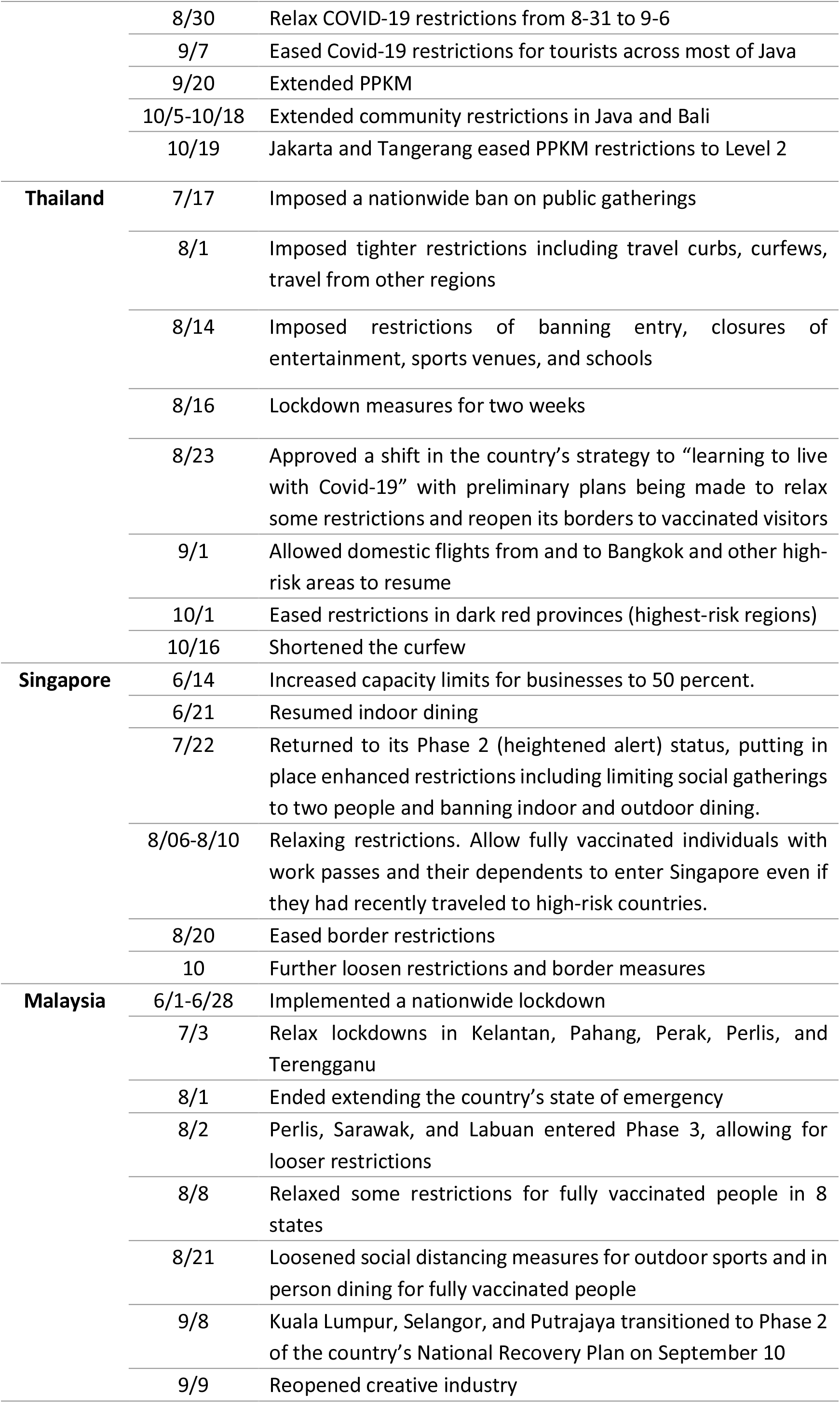
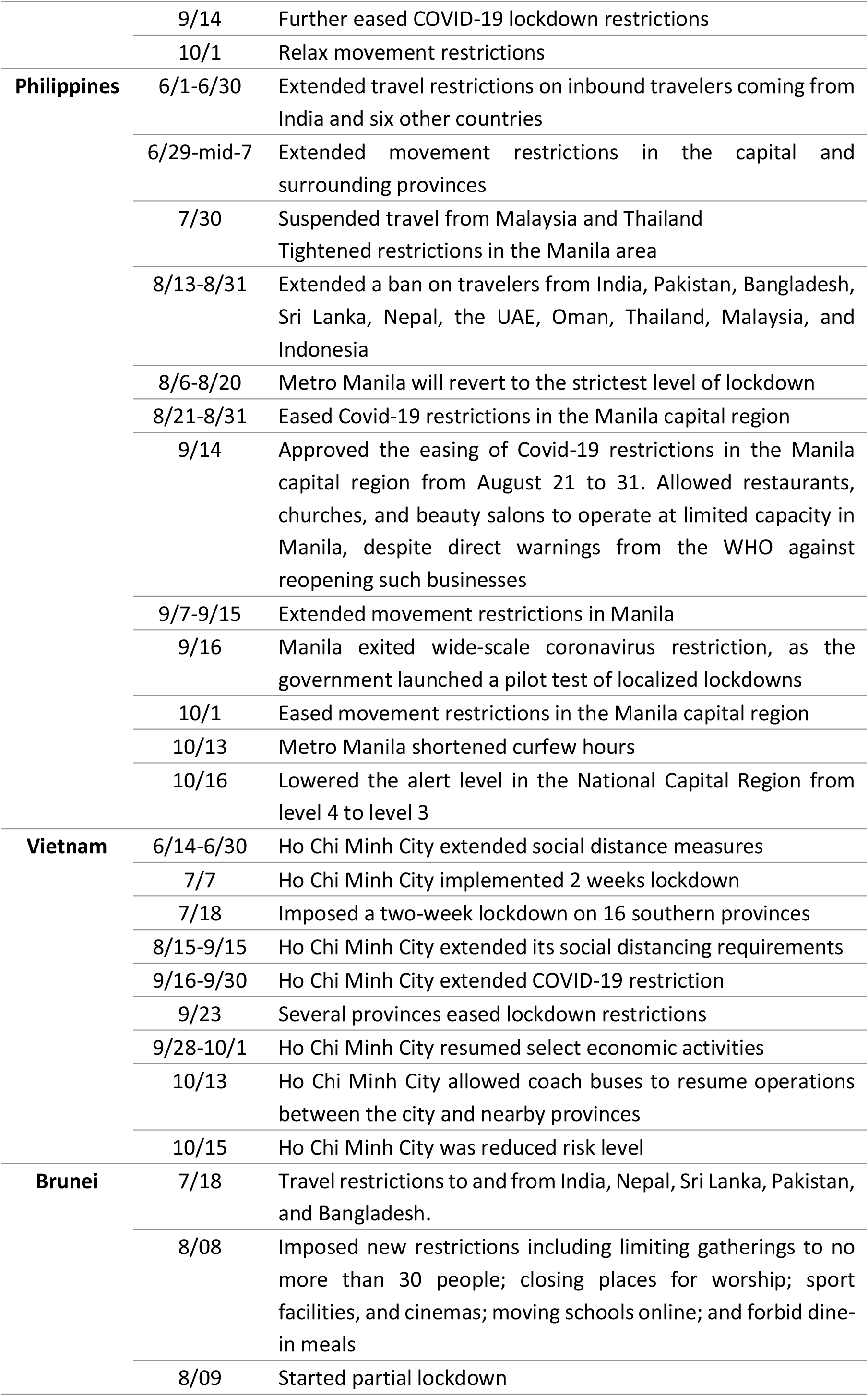
Major public health interventions in SEA.

### 2.2. COVID-19 Daily cases and population

We obtained or extracted data of COVID-19 confirmed cases from the official websites of public health authorities in the seven countries and Johns Hopkins University’s Center for Systems Science and Engineering GIS dashboard (Table 2). We adopted data from 1 June 2021 to 31 October 2021, which is the approximate date of the second COVID-19 outbreak of these seven countries in SEA (Figure 1). We aggregated the data at the first administrative level except for those in Singapore and Brunei which were aggregated at country level considering the similar magnitude of area and population in each analytic unit. We obtained or extracted population data from statistical reports and yearbooks from those countries (Table 2).

**Table 2.**
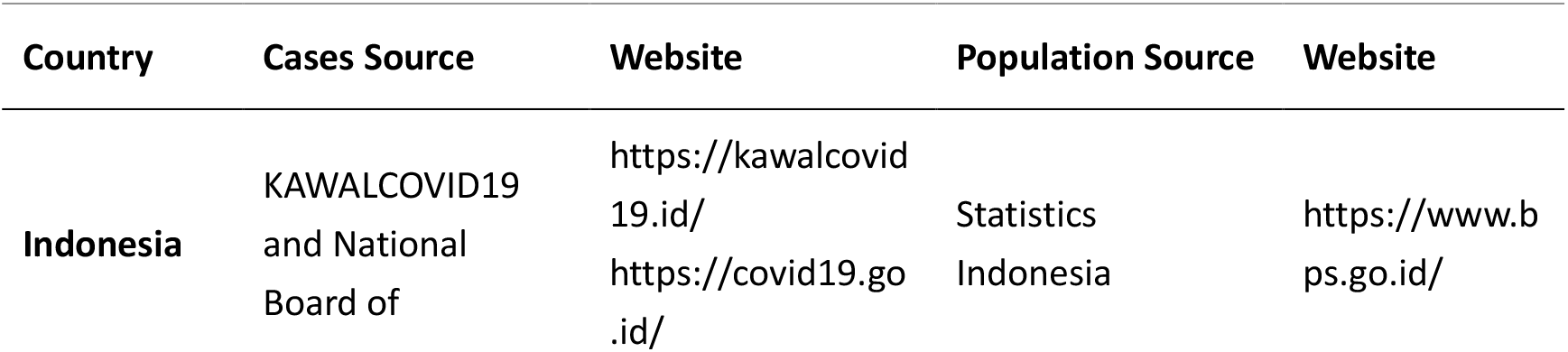
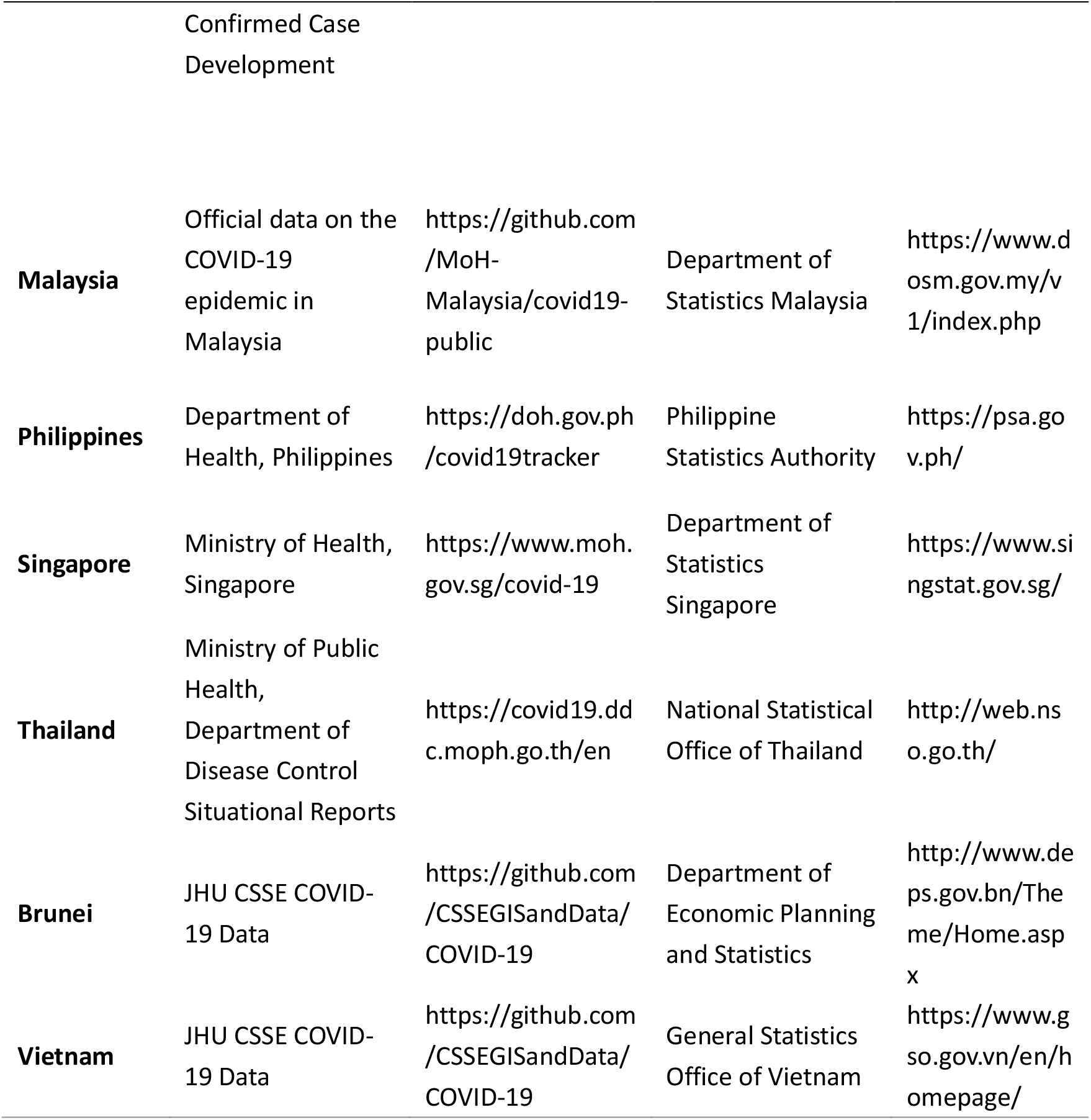
Collected Data and Their Sources.

**Figure 1.**
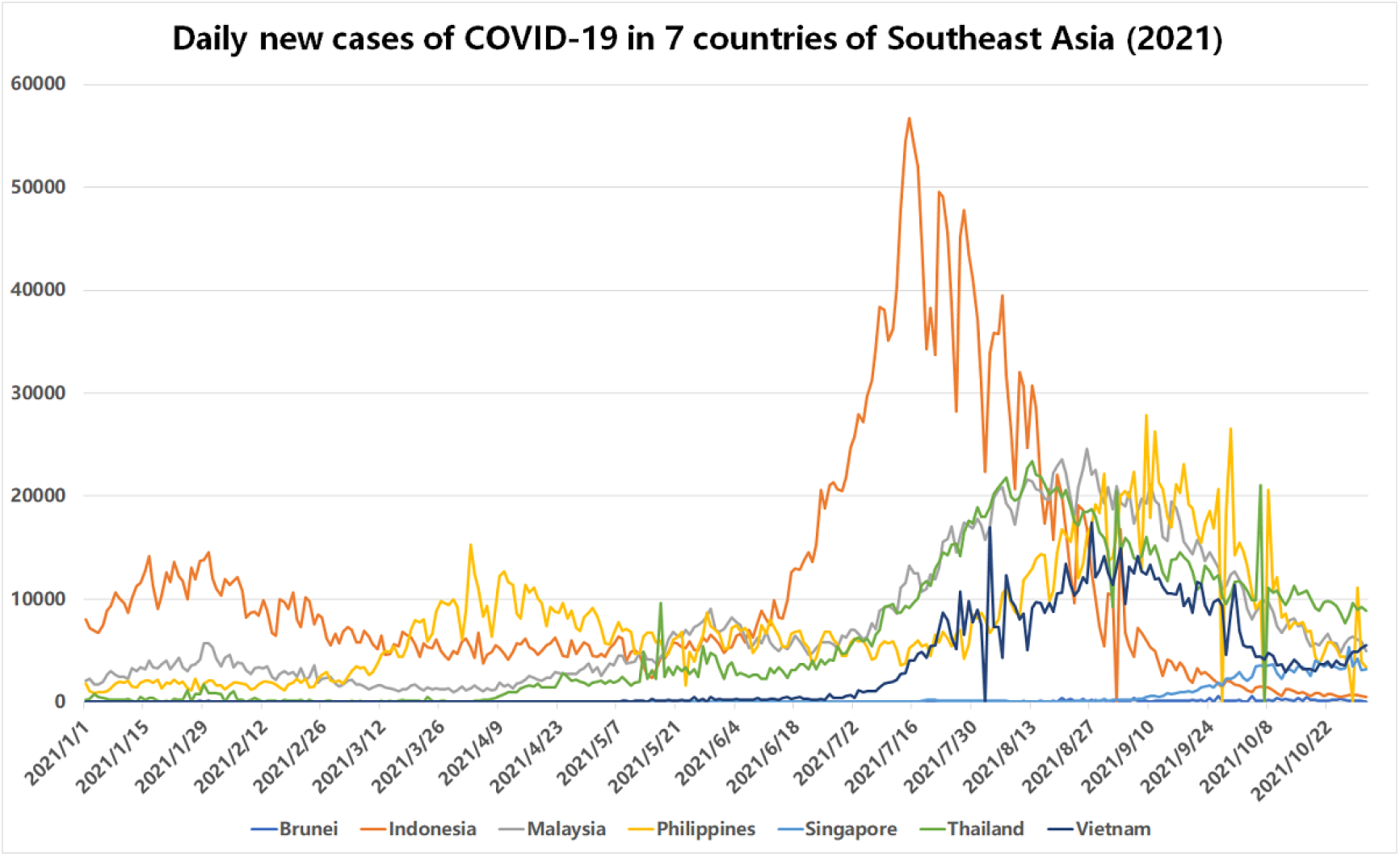
Daily new cases of COVID-19 in SEA (from January 1 to October 31)

**Figure 2.**
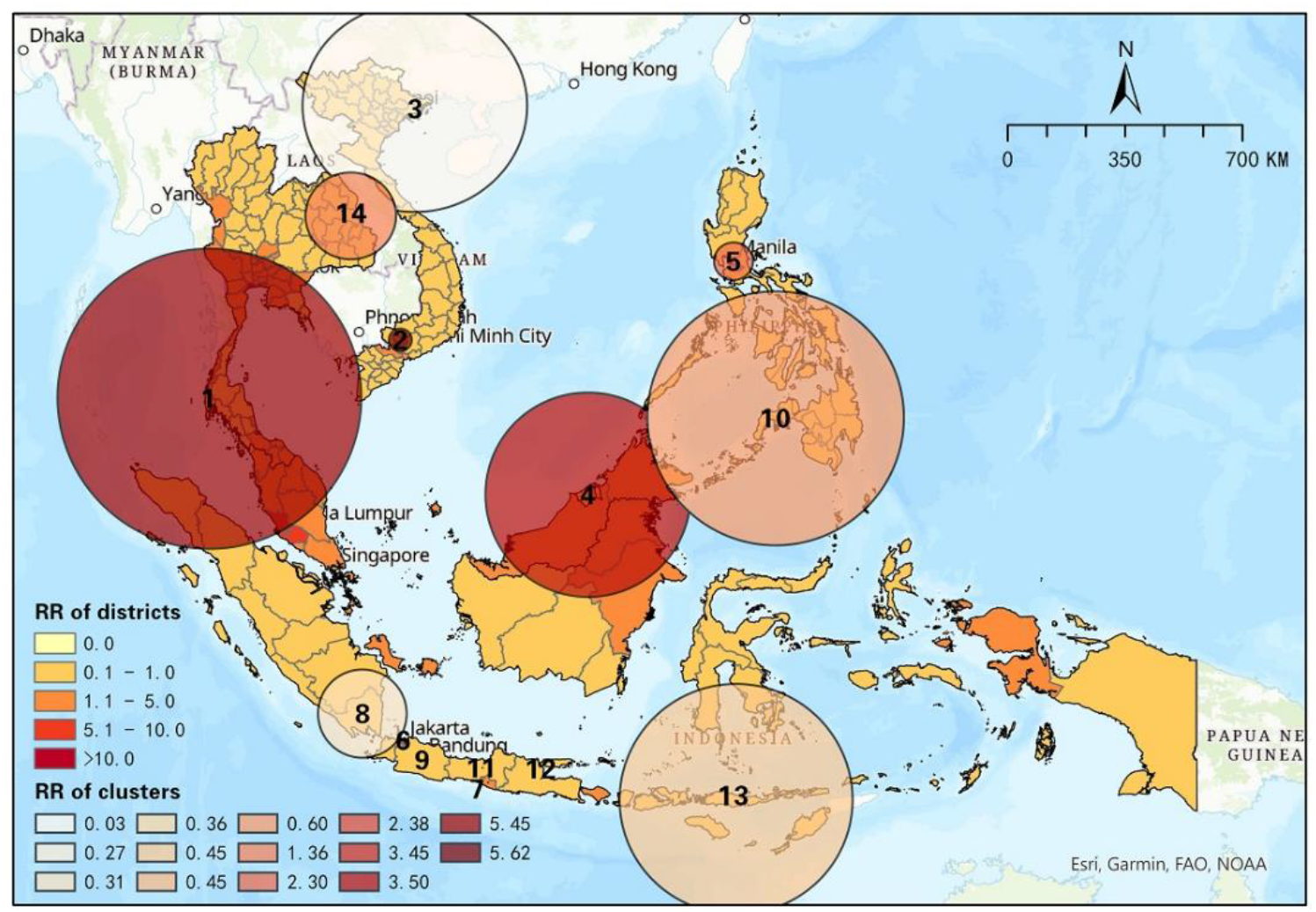
Spatial distribution of space-time clusters of COVID-19 from June 1 to August 31.

### 2.3. Space-Time scan statistical analysis

To explore emerging and active space-time clusters of COVID-19 cases in SEA, we conducted SaTScanTM (version 9.6) [26] to run prospective space-time scan statistic, which is normally used in geographical clusters of epidemic detection [27, 28]. Using space-time scan statistic, we can identify and map significant clusters of the Delta variant in SEA considering the uneven distribution of population size. The space-time scan statistic adopted a cylinder to detect potential space-time clusters in SEA, which can cover each possible location, size, and time period. For each cylinder, the base represents space, the height represents time, and the center represents the centroids of study units throughout SEA. The size of the cylindrical window is expanded until reaching a specific maximum spatial and temporal upper bound which is set to 10% of the population risk and 50% of the study period respectively in this study. Besides, we defined the minimum duration of each cluster to two days for surveillance of the continuously existing clusters. Moreover, the minimum number of cases in each cluster is set to 3 in order to ensure that there must be at least three cases in each cluster.

We assumed that the COVID-19 cases follow a Poisson distribution according to the population of study units in SEA. The null hypothesis H_0_ indicates that the model reflects infection of COVID-19 having a constant intensity μ within or outside the cylinder, which is proportional to at-risk population. Alternative hypothesis H_A_ indicates that the observed cases are more than expected cases, which reflects an increased risk within a cylinder. Expected cases are calculated by Equation (1) [13, 29]:

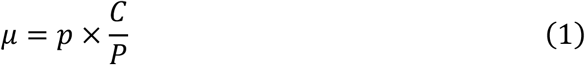

where p represents the population within a study unit, C represents the total COVID-19 cases in our study area (i.e., seven countries in SEA), and P represents the total estimated population within our study area.

A maximum likelihood ratio test is used to evaluate the null and alternative hypotheses. It can identify scanning windows with an elevated risk for COVID-19, which is defined by equation (2) [30, 31]:

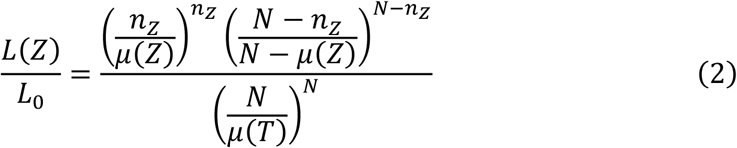

where represents the likelihood function for cylinder Z; represents the likelihood function for H_0_; represents the number of COVID-19 cases in a cylinder; represents the number of expected cases in cylinder Z; N represents the total number of observed cases for the seven countries in SEA across all time periods. When the likelihood ratio is greater than 1, there is an elevated risk in the cylinder, and the cylinder with the maximum likelihood ratio should be the most likely cluster.

The relative risk of COVID-19 is assumed homogeneous throughout different districts within the same cluster. To make the results more reasonable, we calculated relative risk (RR) for each study unit within a cluster to explore the spatial heterogeneity of relative risk of COVID-19 (equation (3)) [32]:

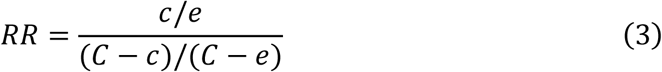

where *c* represents the total number of COVID-19 in a study unit; *e* represents the total number of expected cases in a study unit; *C* represents the total number of observed cases in the seven countries of SEA. The formula indicates that RR represents the estimated risk in a study unit divided by the risk outside that unit. Specifically, if a location (cluster or study unit) has a *RR* of 3, the population within the location are 3 times more likely to be exposed to COVID-19 infection than its outside.

In the following sections, the results reveal significant emerging clusters of COVID-19 cases in seven countries of SEA from 1 June to 31 October. Considering that some SEA countries (e.g., Malaysia, Philippines, Singapore, Thailand) started loosening their restrictions from August along with strengthening their vaccination plan, we divided the timeline into two parts (i.e., June 1 to August 31 and June 1 to October 31) in order to identify the dynamics of clusters. Additionally, we explored the variation of RR across each district in each half-month, which is approximately equal to the incubation period of an infection [33], using a cumulative half a month prospective scanning approach. Then we compared the interventions with the discovered space-time characteristics to identify the potential linkage between political intervention and progression of the Delta variant of COVID-19.

## 3. Results and Discussion

### 3.1. Dynamics of district-level merging clusters in SEA

#### 3.1.1. Results between June 1 and August 31, 2021

As shown in table 3, 14 significant space-time clusters were identified during June 1 and August 31 in SEA, including 7 high-risk clusters (RR > 1) and 7 low-risk clusters (RR < 1). Most of the high-risk clusters emerged between mid-July and late-August, which means the situation of COVID-19 in SEA became severe during this period. Specifically, Cluster 1 was the most likely cluster and a transnational cluster, containing 39 high-risk districts (RR>1) out of 47 districts of Malaysia and Thailand. This cluster had the highest relative risk of 5.45, which means people in this cluster were 5.45 times more likely to be exposed to COVID-19 than other regions. Similarly, cluster 2 had relative risk of 5.62 and contained 2 districts in Vietnam, namely, Binh Duong and Ho Chi Minh city. Another transnational cluster was cluster 4, with an RR of 3.50, containing 6 districts of Malaysia, Indonesia, and Brunei. Additionally, north Thailand and north Philippines also emerged as high-risk clusters from July 22 to August 31, and from August 11 to August 31 respectively. There were also 2 clusters emerging in Indonesia and they contained only 1 district, which was Jakarta, with an RR of 2.38, and Daerah Istimewa Yogyakarta (DIY), with an RR of 3.88. Three high-risk clusters in Indonesia reveal that population within a number of regions in Indonesia were more likely to be exposed to COVID-19 compared to other regions in SEA during this period. Additionally, there were 7 low-risk clusters distributing across other regions of SEA (e.g., north of Vietnam, south of Philippines, some other districts of Indonesia), which means population within these clusters were less likely to be exposed to COVID-19. Note that, cluster 10, with an RR of 0.60, contained 2 high-risk districts in Philippines (i.e., Region VII, with an RR of 1.07, and Region X, with an RR of 1.08). Fig 3 shows the distribution of each cluster. There were a number of small-scale clusters in the south of Indonesia while the largest scale cluster appeared across the south of Thailand and the north of Malaysia. From the results, the Delta variant of COVID-19 had a wider influence in Malaysia and Indonesia in the early phase, while some regions in Vietnam and Philippines had relatively high risk as well.

**Table 3.** Space-time clusters of COVID-19 from June 1 to August 31.

| Cluster | Duration<br>(Days) | No. of<br>Districts | P<br>value | Observed | Expected | RR | No. of<br>Districts<br>(RR >1) |
| --- | --- | --- | --- | --- | --- | --- | --- |
| 1 | 7/17 to<br>8/31 | 47 | <0.001 | 1246176 | 278375.06 | 5.45 | 39 |
| 2 | 7/18 to<br>8/31 | 2 | <0.001 | 300418 | 55874.66 | 5.62 | 2 |
| 3 | 7/17 to<br>8/31 | 30 | <0.001 | 6335 | 223562.03 | 0.03 | 0 |
| 4 | 7/22 to<br>8/31 | 6 | <0.001 | 184097 | 53834.73 | 3.50 | 4 |
| 5 | 8/11 to<br>8/31 | 3 | <0.001 | 196693 | 87355.91 | 2.30 | 3 |
| 6 | 7/17 to<br>8/31 | 1<br>(Jakarta) | <0.001 | 123567 | 52481.33 | 2.38 | 1 |
| 7 | 7/17 to<br>8/31 | 1<br>(DIY) | <0.001 | 62476 | 18229.28 | 3.45 | 1 |
| 8 | 8/7 to 8/31 | 3 | <0.001 | 24861 | 79338.99 | 0.31 | 0 |
| 9 | 8/7 to 8/31 | 1<br>(Jawa Barat) | <0.001 | 59194 | 130362.28 | 0.45 | 0 |
| 10 | 7/17 to<br>8/31 | 11 | <0.001 | 158665 | 259719.07 | 0.60 | 2 |
| 11 | 8/18 to<br>8/31 | 1<br>(Jawa Tengah) | <0.001 | 15132 | 55221.58 | 0.27 | 0 |
| 12 | 8/15 to<br>8/31 | 1<br>(Jawa Timur) | <0.001 | 26939 | 74674.85 | 0.36 | 0 |
| 13 | 8/15 to<br>8/31 | 4 | <0.001 | 18415 | 41030.60 | 0.45 | 0 |
| 14 | 7/22 to<br>8/31 | 14 | <0.001 | 82443 | 60927.09 | 1.36 | 11 |

**Figure 3.**
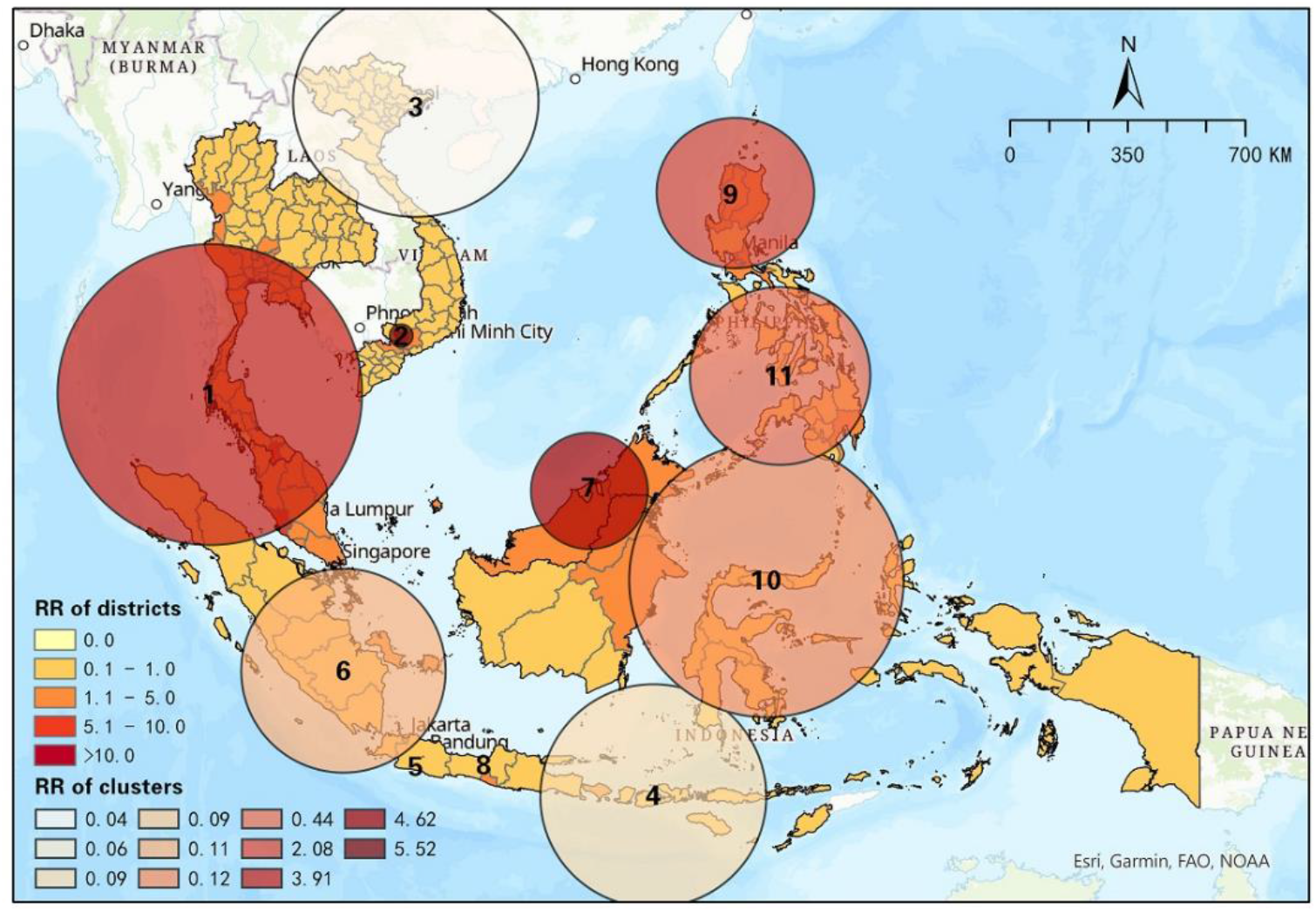
Spatial distribution of space-time clusters of COVID-19 from June 1 to October 31

#### 3.1.2. Results between June 1 and October 31, 2021

Eleven significant clusters were detected between June 1 and October 31, which was three less than the early phase. Among the clusters, there were only 4 high-risk clusters (RR > 1), decreasing from 7 in the previous period (Table 4). The most likely cluster was the same as one of the previous periods, which covered partial districts of Thailand and Malaysia. The RR of cluster 1, however, decreased from 5.45 to 3.91 in this period, while the number of high-risk districts increased from 39 to 45, which implies that more districts were influenced by COVID-19. Cluster 2 was also the same as one of the previous periods, containing Binh Duong and Ho Chi Minh city. Besides, cluster 7 evolved from cluster 4 in the previous period, with the exclusion of Kalimantan Timur. The RR of this cluster increased from 3.50 to 4.62, which means that people in this cluster were more likely to be infected in this period. Similarly, Cordillera Administrative Region, Region II, and Region I merged with one high-risk cluster in the north of Philippines and formed a larger cluster. The rest of the clusters were low-risk clusters with RR less than 1. Figure 3 visualizes the distribution of clusters in this period, which shows directly that some of the clusters remained between 2 continuous periods, while a number of clusters in the previous period disappeared and some new clusters appeared in this period. Especially in Indonesia, high-risk clusters in Jakarta and DIY disappeared, which emerged low-risk clusters with other districts. Overall, the space-time scan statistic results show the transmission and dispersal of the Delta variant of COVID-19 in SEA from 2 different time periods.

**Table 4.**
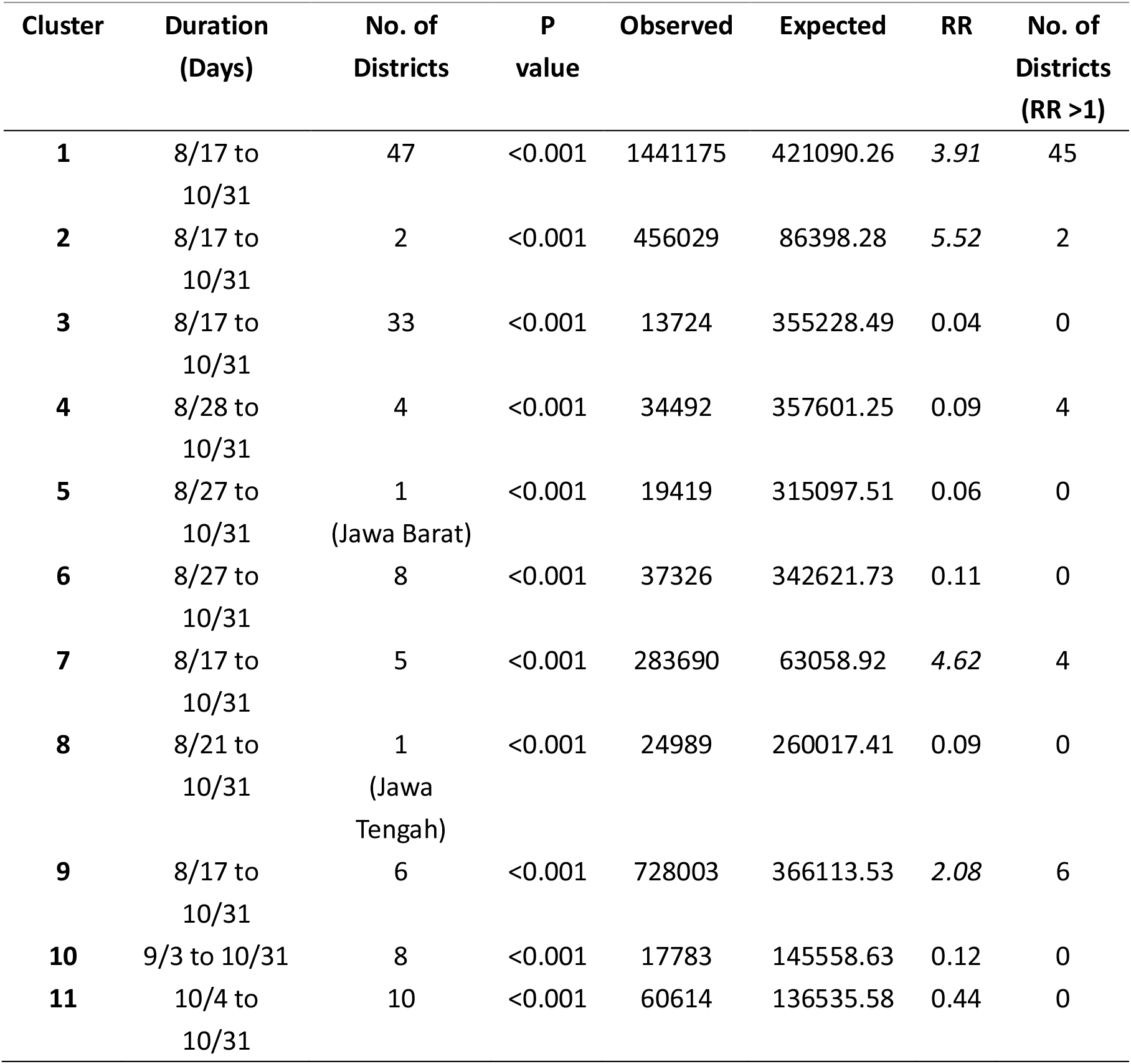
Space-time clusters of COVID-19 from June 1 to October 31.

| Cluster | Duration<br>(Days) | No. of<br>Districts | P<br>value | Observed | Expected | RR | No. of<br>Districts<br>(RR >1) |
| --- | --- | --- | --- | --- | --- | --- | --- |
| 1 | 8/17 to<br>10/31 | 47 | <0.001 | 1441175 | 421090.26 | 3.91 | 45 |
| 2 | 8/17 to<br>10/31 | 2 | <0.001 | 456029 | 86398.28 | 5.52 | 2 |
| 3 | 8/17 to<br>10/31 | 33 | <0.001 | 13724 | 355228.49 | 0.04 | 0 |
| 4 | 8/28 to<br>10/31 | 4 | <0.001 | 34492 | 357601.25 | 0.09 | 4 |
| 5 | 8/27 to<br>10/31 | 1<br>(Jawa Barat) | <0.001 | 19419 | 315097.51 | 0.06 | 0 |
| 6 | 8/27 to<br>10/31 | 8 | <0.001 | 37326 | 342621.73 | 0.11 | 0 |
| 7 | 8/17 to<br>10/31 | 5 | <0.001 | 283690 | 63058.92 | 4.62 | 4 |
| 8 | 8/21 to<br>10/31 | 1<br>(Jawa<br>Tengah) | <0.001 | 24989 | 260017.41 | 0.09 | 0 |
| 9 | 8/17 to<br>10/31 | 6 | <0.001 | 728003 | 366113.53 | 2.08 | 6 |
| 10 | 9/3 to 10/31 | 8 | <0.001 | 17783 | 145558.63 | 0.12 | 0 |
| 11 | 10/4 to<br>10/31 | 10 | <0.001 | 60614 | 136535.58 | 0.44 | 0 |

### 3.2. Temporal progression of relative risk of COVID-19 in SEA

Figure 4 presents the changes of District RR of COVID-19 in SEA between two outbreak periods, 1 June to 31 August and 1 June to 31 October. Overall, this temporal change in RR manifests different space-time characteristics of the progression of COVID-19 in different districts before and after countries changed their intervention strategies in handling COVID-19 in July and August. Amidst the seven SEA countries, Indonesia was the only country that showed an overall positive trend of decreasing RR in every district, while alarming changes of increasing RR were frequently seen in other six countries especially Singapore, Philippines, Malaysia and Vietnam.

**Figure 4.**
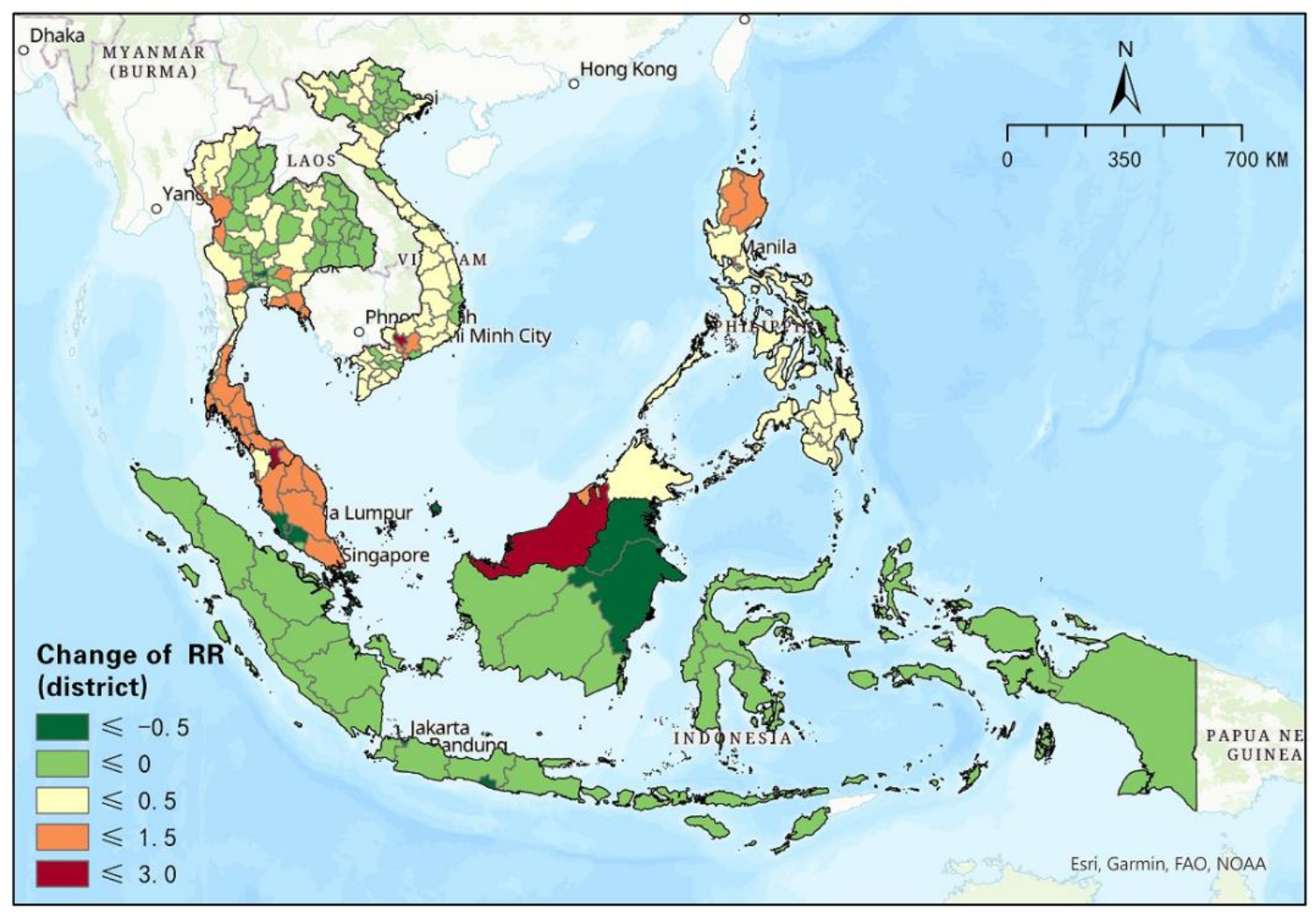
Changes in relative risk of COVID-19 (district level) between two periods (June 1 to August 31 and June 1 to October 31)

Specifically, all districts of Indonesia observed a decrease in RR to different extents. While RR in most districts of Indonesia decreased slightly between 0.05 to 0.5, the other five districts (i.e., Jakarta, Daerah Istimewa Yogyakarta, Kalimantan Utara, Kalimantan Timur, and Kepulauan Riau) manifested a rather significant decrease (≤-0.5), in which the highest difference (−1.4) between the two phases was seen in Jakarta, the capital of Indonesia. Note that Jakarta was one of the major emerging risk districts early in the second outbreak (Figure 5) and was still faced with a relatively high RR (2.79) until the end of our study period. Nevertheless, characterized as the largest city nationwide with a high-density population, Jakarta’s success in preventing exacerbation of risk impact of COVID-19 is most likely to be related to its persistent restrictions and unchanged strategy towards COVID-19. Ever since June 1, all provinces in Indonesia have implemented Micro Community Activity Restrictions (PPKM) which includes policies of mandatory work from home, guidance on online teaching and restrictions of dine-in, social gathering and inter-province as well as international traveling [34]. On the contrary, RR of all districts in the Philippines have increased between the two outbreak periods, indicating an overall deterioration in the risk impact of COVID-19. Fortunately, amidst a total of 17 districts in the Philippines, 14 of which showed minor increases (≤0.5). Other three districts, namely National Capital Region, Region II and Cordillera Administrative Region manifested an increase between 0.59 to 1.23. Meanwhile, no significant increase (>1.5) was observed, indicating that the most severe variation of RR did not occur in the Philippines. This is mainly because Philippines loosened restrictions on a wide scale, including reopening restaurants and churches, as well as replacing a larger scale of coronavirus restrictions with localized lockdowns, though it did take certain measures to intervene in the epidemic outbreak, such as the strictest level of lockdown in Metro Manila and online teaching which may have helped prevent significant increases in RR. The gradually intensified risks of districts in the Philippines should receive continuous attention.

**Figure 5.**
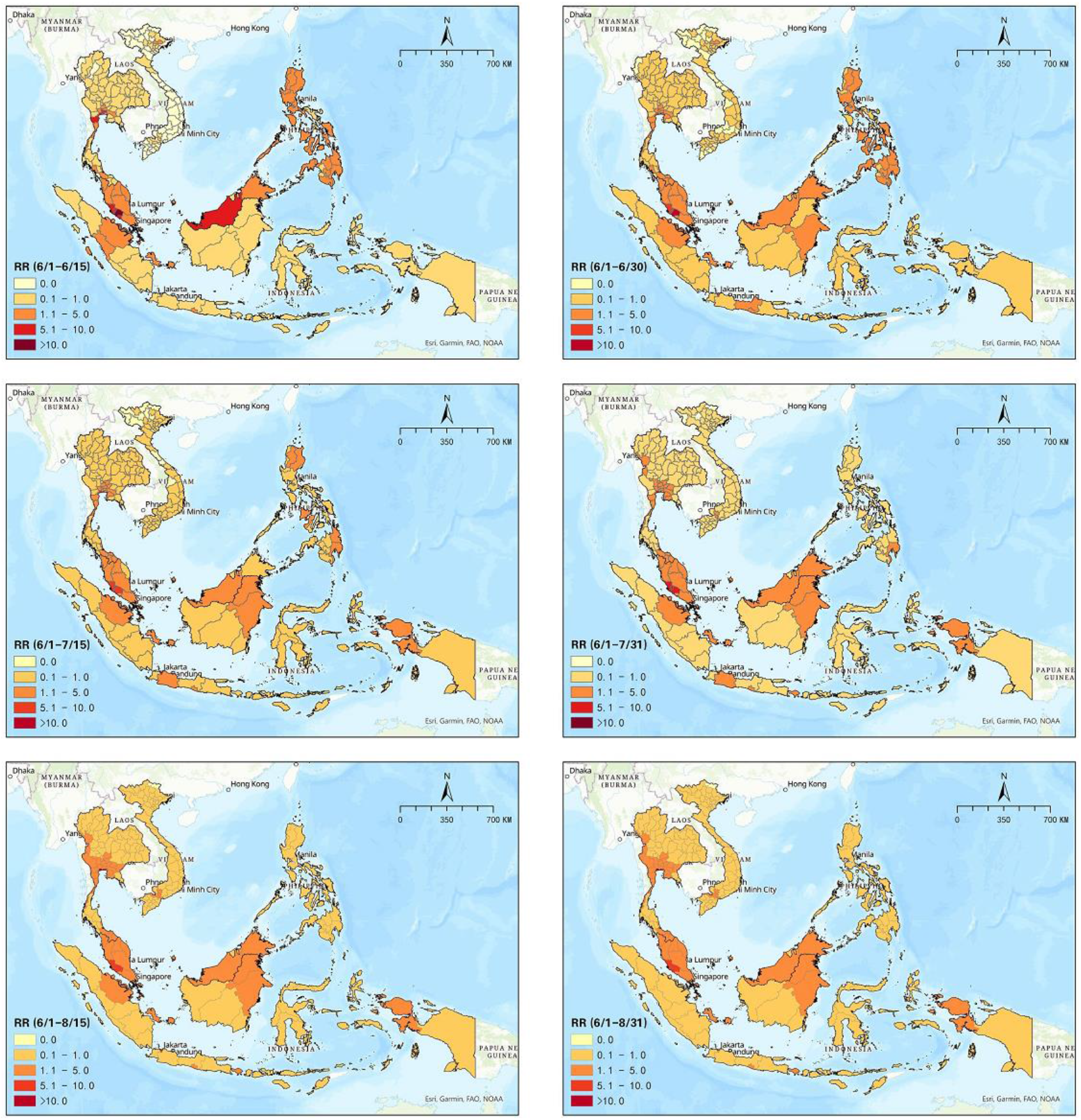
Spatial patterns of progression of COVID-19 relative risk in SEA (June 1 to June 15, June 1 to June 30, June 1 to July 15, June 1 to July 31, June 1 to August 15, June 1 to August 31)

Other countries (Singapore, Malaysia, Brunei, Thailand, Vietnam), on the other hand, showed a variety of RR changes in different districts. Approximately half of which (39) showed increases in RR, and half decreased amid the 77 districts in Thailand. It is obvious that coastal areas in the south of the country were faced with more elevated RR than inland areas in the central, eastern and northern districts. RR in almost all districts in Vietnam changed slightly, ranging from -0.08 to 0.28 except for 3 connected cities (i.e., Dong Nai:0.63, Ho Chi Minh: 0.72 and Binh Duong:1.73). Particularly, although a series of regulations and restrictions including lockdown, curfew, social distancing measures etc., had been specifically implemented in Ho Chi Minh, the city still manifested overall worsen symptoms. This may be related to the high contagiousness of Delta variant and overloaded health-care system in Ho Chi Minh City [35]. Also, Ho Chi Minh City has a larger population base and more developed economy than other districts which resulted in more infections via contact with a relatively larger number of crowds. It has been suggested that population density and contact intensity are the main drivers for the propagation and amplification of this virus [36]. As for Malaysia, 10 out of 16 districts showed an increase in RR and took up a major proportion of the country. Although Labuan and Kuala Lumpur observed the most obvious decrease among SEA districts, it should be noted that they were also the most severely threatened districts by COVID-19 risks early in the study period (RR>10) and remain the relatively highest RR amidst SEA until the end of the study. This is probably because the Malaysian government gradually adopted more loose measures than the other SEA countries with its phase development of the National Recovery Plan since the beginning of our study period. Additionally, Singapore saw an increased RR of 1.43, which is probably related to its intermittent relaxation of social restrictions. RR in Brunei also increased, but it still remained less than 1.

Furthermore, we detected the progression of RR at half a month interval, and 10 intervals were used to illustrate the progression of the Delta variant from June to October (Figure 5, Figure 6). In the first half of June, elevated risks were identified in the middle south of Thailand, many states in Malaysia, and many districts in Philippines, while 44 out of 63 districts in Vietnam manifested RR=0 during this period (Figure 5). These reveal that those high-risk districts were more likely to be in danger of a new wave of epidemic. From then on, the Delta variant spread in SEA, and the capital areas of several countries have been severely affected in SEA (e.g., Bangkok Metropolis and surrounding areas, Kuala Lumpur and surrounding areas, Jakarta and surrounding areas, Manila capital areas, as well as Ho Chi Minh city). By the middle of August, Thailand had been influenced by the expansion of COVID-19, and most districts showed an elevated risk. On the contrary, RR in the Manila capital region in Philippines decreased, which was probably due to suspension of travel from Malaysia, tightened restrictions, and curfew extension in late July and early August. Additionally, most high-risk areas in the previous months kept severe in this time, although the relative risk of some districts slightly declined (e.g., Bangkok, Samut Sakhon, Jakarta, Riau, Sarawak, etc.). Note that, most of the countries tightened the restriction before this time except for Malaysia, which loosened their restrictions. This phenomenon also reflected the strong infection and danger of the Delta variant. In the next one and a half month (i.e., August 15 to September 30), the situation of Indonesia had been improving, which was inseparable with continuous extension of activity restrictions in Indonesia (e.g., extended PPKM for the ninth time). The north of Philippines, however, showed increased relative risk in mid-September, which may be due to the relaxing restriction of Manila capital region on August 19 and mid-September (Figure 6). In the next month, relative risk of Singapore increased from 0.44 to 1.53, which was probably due to gradually loosened restrictions. Besides, the situation across north of Malaysia and south of Thailand was getting worse as well. Up to October 31, the situation of Thailand, Malaysia, Singapore, capital areas in Vietnam, and capital areas in Philippines was still alarming, which should be noticed with further surveillance by obtaining daily case reports in the future.

**Figure 6.**
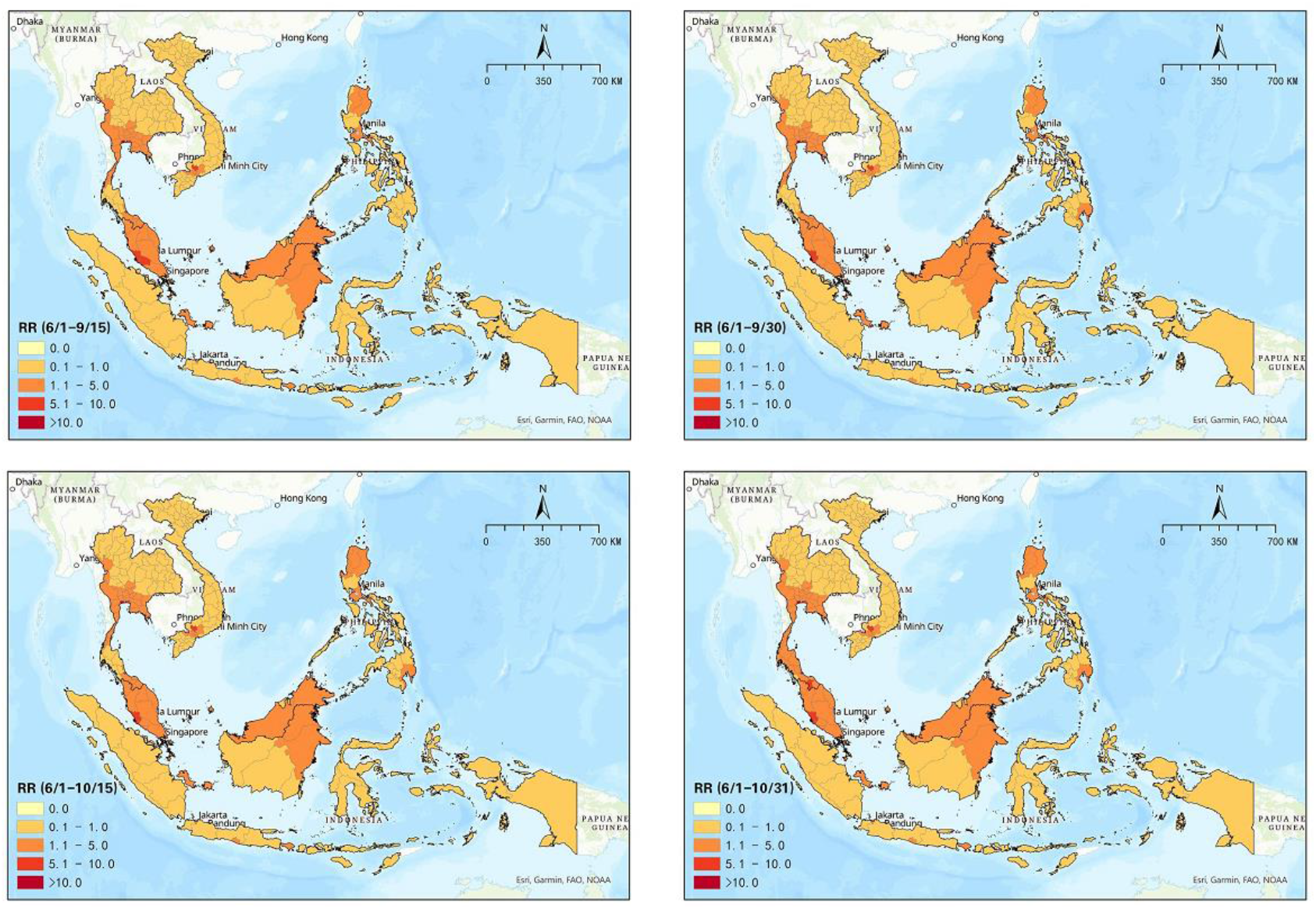
Spatial patterns of progression of COVID-19 relative risk in SEA (June 1 to September 15, June 1 to September 30, June 1 to October 15, June 1 to October 31)

## 4. Discussion

### Principal Findings

In this study, we utilized prospective space-time scan statistics to detect the emerging and existing space-time clusters of COVID-19 in SEA. We found that most districts in Malaysia and Philippines, the capital and its surrounding areas in Thailand, Vietnam, and Indonesia exhibited high risk of COVID-19 transmission in the early phase (June to August 2021) during our study period. Space-time clusters and relative risk of districts changed along with the dynamics of policies introduced by each country after August. Indonesia successfully mitigated the risk of this epidemic by implementing continuous restrictions, while a number of regions in Malaysia, Singapore, Thailand, and Philippines remained at a relatively high risk of COVID-19 transmission due to diverse degrees of relaxation. Although propagation of COVID-19 was influenced by diverse factors (e.g., social economy, population, resource allocation, etc.), continuous strict restrictions were beneficial on epidemic control, especially for those developing regions with weak public health systems and relatively low vaccination rates. To our knowledge, this is the first attempt to explore the space-time progression of delta variant outbreak in SEA, as well as summarize the potential linkage between the epidemic dynamics and diverse public health policies. Spatio-temporal statistics is an effective method to identify high-risk clusters and regions [37], which can help explore spatial temporal patterns of epidemic progression and better understand the dynamics and characteristics of the Delta variant in SEA. Furthermore, by combining the results with relevant measures that reflect the appropriateness of public health interventions, governments can also timely improve the interventions. In the meantime, health authorities can timely adjust their interventions by comparing the RR with current measures, which may reflect the appropriateness of these interventions. In short, the prospective space-time scan can continuously be used to monitor the dynamics of COVID-19 with the most updated data, and timely adjust the potential gaps in domestic public health intervention to prevent further deterioration of the pandemic.

### Implications and Recommendations

Public health interventions played an important role in epidemic containment, in which social restriction policies effectively mitigate the propagation of COVID-19 [29]. Restrictions of mass gathering and travel, keeping social distance, and reducing human mobility are beneficial to control COVID-19 because these measures could reduce the probability of exposure to virus infection [38, 39]. Our study discovered the potential linkage between the dynamics of outbreaks and interventions. This indicates that while continuously implemented strict restrictions can prevent exacerbation of the pandemic, temporary or continuous relaxation may result in an acceleration of the epidemic propagation. Appropriate restriction policies are key to preventing the pandemic because high transmission of the COVID-19 variant can easily lead to worse situations. Also, if the number of community cases exceeded imported cases, border restriction would be less valuable than domestic measures. In this case, authorities should emphasize more on domestic intervention in order to reduce community transmission [40]. Nevertheless, it is well known that those intervention measures against COVID-19 come with economic costs from other perspectives (i.e., labor market etc.). When making intervention regulations, economic and social justifications for public health policies are one of the priorities that governments would consider [41]. Hence, it is one of the challenges for all countries to live with COVID-19 and weigh the balance between epidemic development and social economic loss [42]. Given the social and economic status, most countries in SEA have decided to change their strategies from the elimination of cases to living with COVID-19 since August 2021 despite the new outbreak of Delta variant [8]. A concurrent trend observed in SEA is that all countries except for Indonesia have been gradually loosening social restrictions, allowing international communications whilst boosting vaccination attempting to achieve group immunity among citizens. For instance, the Singapore Ministry of Health believes that with the assistance of a high vaccination rate, it is assured that the growth of COVID-19 cases, though cannot be eliminated, will be controlled at a certain level. By doing this, economy and social norms could be restored without causing another disease outbreak or a breakdown in the hospital system [43]. Vaccination is increasingly essential to protect the crowd from the exacerbating threat of morbidity and mortality of this Delta variant and future variant [44]. Effective vaccination is highly beneficial to avert infection, while wide-scale vaccination is proven to be able to successfully control transmission of COVID-19 so far [45-47]. However, it should be noted that vaccinations cannot be depended on entirely. The previous experiment found that effectiveness of available vaccines against the Delta variant (B.1.617.2) showed a reduction compared to previous virus variants [48], implying that current vaccination is highly likely to become ineffective in defending against many mutations in the future [49]. On the other hand, considering the fragile health systems in SEA, contact tracing, quick isolation and strict restrictions are still essential to avoid any miscalculation and prevent another outbreak [50, 51].

International coordination including vaccine allocation, food, finance, and equitable access is essential for the world to respond to the pandemic [52]. The pandemic did not only cause the breakdown of developing countries, but also developed countries, which indicates that countries should enhance transparency, share information, and strengthen cooperation to protect international health and security [53]. As for Southeast Asia, due to strong socioeconomic connectivity among all countries, it is vital for countries in SEA to adopt collaborative policies to overcome the challenges of COVID-19. Sustainable development in SEA during the pandemic era requires multi-sectoral cooperation including tourism, economy, health etc., which can help effectively coordinate border restrictions, international trade, or allocation of medical supplies. To ensure cooperation among countries in SEA, regional surveillance and information sharing should be indispensable for each country, which can support more precise control and prevention of COVID-19. Although some countries (Singapore, Vietnam) have made efforts to donate medical equipment and provide aid to their neighbors [3], we hope that there will be more interactive assistance among countries in SEA in the future, especially along the continuous emergence of COVID-19 variants. Furthermore, multilateral collaboration with countries or regions (e.g., European Union) outside SEA is considered as an effective and valuable way to help respond to the pandemic, although there will still be a long way to go in the future [3]. Overall, SEA should focus on regional coordination with long-term plans to solve a series of threats caused by the epidemic, considering the long-term challenges caused by the COVID-19 pandemic.

### Limitations and Future Work

Despite the insights from our study, there are notable limitations in the COVID-19 data. There are only seven out of twelve countries provided with data at the primary administrative district level, so we were not able to explore the complete propagation process in SEA. In addition, if high spatial resolution data become available (i.e., city, county, even block or subzone), more specific and detailed patterns could be revealed.

As a matter of fact, many previous studies were also faced with a shortage or loss of available data (i.e., insufficient pediatrics data) [54-56]. Moreover, insufficient knowledge of the data or dynamics would lead to invalidity and unreliability of responses to COVID-19 [57]. Therefore, we strongly suggest that public health authorities should disclose more representative and reliable data [58, 59]. Apart from this, COVID-19 transmission and its impacts have shown environmental inequality in terms of household income, education level, age, gender etc. [60, 61]. The potential correlation between environmental inequality and COVID-19 should be further studied using diverse data, which can provide significant insights to resource allocation and regional prevention.

## Data Availability

All data produced in the present study are available upon reasonable request to the authors

## Notes

### Competing Interest Statement

The authors have declared no competing interest.

### Funding Statement

This study did not receive any funding

